# Early life adversity increases risk for chronic posttraumatic pain, data from humans and rodents

**DOI:** 10.1101/2024.11.01.24316303

**Authors:** Lauren A. McKibben, Alice Woolard, Samuel A. McLean, Ying Zhao, Taanvii Verma, Jacqueline Mickelson, Hongxia Lu, Jarred Lobo, Stacey L. House, Francesca L. Beaudoin, Xinming An, Jennifer S. Stevens, Thomas C. Neylan, Tanja Jovanovic, Laura T. Germine, Scott L. Rauch, John P. Haran, Alan B. Storrow, Christopher Lewandowski, Phyllis L. Hendry, Sophia Sheikh, Christopher W. Jones, Brittany E. Punches, Lauren A. Hudak, Jose L. Pascual, Mark J. Seamon, Claire Pearson, David A. Peak, Roland C. Merchant, Robert M. Domeier, Niels K. Rathlev, Brian J. O’Neil, Leon D. Sanchez, Steven E. Bruce, John F. Sheridan, Ronald C. Kessler, Karestan C. Koenen, Kerry J. Ressler, Sarah D. Linnstaedt

## Abstract

Traumatic stress exposures (TSE) are common in life. While most individuals recover following a TSE, a substantial subset develop adverse posttraumatic neuropsychiatric sequelae such as chronic posttraumatic musculoskeletal pain (CPMP). Vulnerability factors for CPMP are poorly understood, which hinders identification of high-risk individuals for targeted interventions. One known vulnerability factor for many pain types is exposure to early life adversity (ELA), but few studies have assessed whether ELA increases risk for CPMP. This study used data from the AURORA study, a prospective human cohort study of TSE survivors, to test the hypothesis that ELA increases risk for CPMP. In addition, in secondary analyses, we assessed which subtypes of ELA (including childhood bullying) were most predictive of CPMP and whether a rat ELA model consisting of neonatal limited bedding (NLB), combined with single prolonged stress (SPS) in adulthood, would accurately model human findings. In AURORA study participants (n=2,480), using multinomial logistic regression modeling of four identified latent pain classes, we found that ELA increased vulnerability to the high unremitting pain class (OR=1.047, *p*<0.001), the moderate pain class (OR=1.031, *p*<0.001), and the moderate recovery pain class (OR=1.018, *p*=0.004), with physical abuse, emotional abuse, and bullying being the strongest predictors of high pain class assignment. Similarly, in male and female Sprague Dawley rats, in comparison to SPS alone NLB combined with SPS caused increased baseline sensitivity and prolonged mechanical hypersensitivity (F(11,197)=3.22, *p*<0.001). Further studies in animals and humans are needed to understand mechanisms by which ELA confers vulnerability to CPMP.

**Summary:** In humans and rats, early life adversity is associated with a greater duration of musculoskeletal pain and mechanical hypersensitivity following traumatic stress exposures during adulthood.

## INTRODUCTION

Traumatic stress exposures (TSE) affect >90% of individuals in their lifetime.[64] Most individuals recover following TSE but a substantial subset develop chronic post-traumatic musculoskeletal pain (CPMP).[41; 61; 76] CPMP commonly develops independent of tissue injury and is co-occurring with neuropsychiatric outcomes of TSE including posttraumatic stress and depressive symptoms.[6; 7; 39; 50] In addition, CPMP is associated with increased individual and societal burdens including interference with daily activities[6; 39; 75], increased risk for substance and opioid misuse[59], and substantial health care costs[29]. Biologic vulnerability factors and mechanisms of CPMP development remain poorly understood.

Increasing evidence suggests that individuals with a history of early life adversity (ELA) are particularly vulnerable to CPMP. Unfortunately, ELA, such as childhood abuse, neglect, and bullying, are common, affecting approximately half of all children worldwide[30; 40]. Individuals who have experienced considerable ELA are at substantially increased risk of adverse neuropsychiatric outcomes that often co-occur with CPMP.[2; 21; 37; 94] Further, ELA has been shown to increase the severity and chronicity of a variety of acute and chronic pain states[3; 13; 17; 27; 37; 45; 54; 70; 78; 81; 84; 90; 91], including multisite chronic pain[54], fibromyalgia[3], migraine[78], irritable bowel syndrome[45], and chronic fatigue syndrome[37]. Pain-like hypersensitivity has been tested in several different rodent models of ELA, with most studies assessing the effects of either maternal separation or neonatal limited bedding (NLB) models on hypersensitivity (Reviewed in [15]). These rodent models have been associated with increased sensitivity to various reflexive[19; 31; 32; 48; 66; 80], inflammatory[32; 80], and visceral[19; 48; 66] pain-like behaviors[31; 66]. However, previous research assessing the relationship between ELA and CPMP using parallel translational studies in humans and animals is lacking.

In the current study, we used longitudinal outcome data from 2,480 adult multiethnic TSE survivors enrolled in the AURORA study[62] to evaluate the hypothesis that ELA increases risk for CPMP. Specifically, we examined the association between childhood trauma and bullying and longitudinal CPMP course (CPMP latent class trajectory) using multinomial regression analyses. In addition, in secondary analyses we compared the influence of different types of ELA (e.g., emotional versus physical abuse) on longitudinal CPMP course. Finally, we repurposed two animal stress models, the NLB model and the single prolonged stress (SPS) model to assess whether such a combined model could be used in future translational studies as a model of human ELA and CPMP.

## METHODS

### Human Studies

#### The AURORA Cohort

Self-report data was collected as a part of a national, multisite, emergency department-based study of adverse traumatic stress outcomes: Advancing Understanding of RecOvery afteR traumA (AURORA)[62]. AURORA was approved by the Institutional Review Board at all participating institutions. Patients who reported to one of 29 US emergency departments (EDs) within 72 hours of a traumatic stress exposure between September 2017-December 2020 were recruited to participate. Basic study inclusion criteria were English comprehension, ability to understand and agree to the enrollment procedure, hospital admission for no more than 24 hours if at all, and no significant physical injury requiring hospitalization. All participants included in the current study (n=2,480, (“Freeze 4” of the AURORA dataset)) provided written informed consent prior to completing their initial assessment in the ED. Data relevant to the current study were collected in the ED and at follow-up visits 2-weeks following the index TSE for which they were enrolled. Longitudinal pain data was collected using flash surveys on participants’ cellphones. Additional details on the AURORA study have been published[62].

### Study Measures

#### Childhood Trauma Questionnaire

At the 2-week follow-up timepoint, AURORA participants responded to eleven questions from an abbreviated Childhood Trauma Questionnaire[9] which were selected to assess the participant’s experience with five subtypes of ELA: physical abuse, emotional abuse, sexual abuse, physical neglect, and emotional neglect. Responses to each question were rated on a 5-point Likert scale from never (0) to very often (4). As a part of the AURORA study design, the number and type of survey questions administered were carefully selected to minimize response fatigue[24] and collect high quality data across all dimensions assessed. Thus, the CTQ[9] or CTQ-short form[10] were not administered in their entirety. Additional details on CTQ in the context of the AURORA study have been previously reported.[89; 94] Analyses presented here used either a composite CTQ score that added scores from each subscale or CTQ subscores separately. Each CTQ subscore was calculated by summing the score from the 2-3 questions corresponding to each of the five ELA subtypes (sexual abuse subscore range: 0-12, all other subscores range: 0-8, composite score range: 0-44). For dichotomous ELA subtype analyses, a score ≥1 was used as the cutoff for having experienced any of that ELA subtype. Following identification of the top ELA subtype predictors of CPMP, we tested a model using a new ELA composite score based only on the sum of the top three most predictive susbscores.

#### Bullying

Childhood history of bullying was included as an additional measure of ELA, complementary to the CTQ that likely reflects adverse peer-centered experiences, that often occur outside of the home.[11] Bullying measured at the 2-week follow-up timepoint using two questions from the structured clinical interview for the DSM-IV[28] : “how often did other kids call you names or say mean things” and “how often did other kids threaten to hurt you”. Participants responded on a 0-4 Likert scale and the scores for these two questions were combined to create a composite bullying variable with score ranging from 0-8.

#### Latent Pain Class Trajectories

In a previous publication using a subset of the AURORA dataset[72], cellphone-administered flash survey pain data was used to develop latent pain trajectory groups. These flash surveys were administered to participants on days 1, 9, 21, 31, 43, and 53 following their initial TSE. At each time, participants were asked to rate their worst pain and average pain in the last 24 hours using a 0-10 numeric rating scale (NRS). Briefly, latent growth curve models and growth mixture modeling of all patients in the present sample established four pain trajectory groups. Based on longitudinal trajectories these were termed low pain, moderate recovery, moderate pain, and high pain[6; 72]. Predicted membership in one of these four latent class groups was used as the outcome variable for all analyses.

#### Covariates

Our analyses controlled for socio-demographic variables assessed at the ED: sex, age, race/ethnicity, BMI, and Area Deprivation Index (ADI). Sex was assessed using the question “what was your gender as written on your birth certificate?” Participants selected the most appropriate race/ethnicity from the options “Hispanic”, “non-Hispanic white”, “non-Hispanic black”, and “other”. ADI was assessed as previously reported using an online tool (version 3.1, 2019)[87]. ADI summarizes, in a single variable, metrics of income, education, employment, and housing quality based on residential address. An ADI score of 100 indicates the highest level of disadvantage compared to other locations in the US.

Additional covariates were ED/trauma-related variables including the enrolling ED site, trauma type, and Peritraumatic Distress Inventory[14] (PDI) as well as past pain. ED sites were categorized by US region and included “Northeast”, “Southeast”, and “Midwest” (termed “ED region”). Trauma type was characterized by three categories including “Motor Vehicle Collision (MVC)”, “Physical/Sexual Assault”, and “Other”. PDI was assessed using the 8-item Peritraumatic Distress Inventory. Pain experienced in the 30 days before TSE was assessed in the ED using an 11-point, 0-10 NRS (0-no pain to 10-maximum possible pain).[25; 26; 47] The pre-pain variable was binary coded indicating presence or absence of NRS ≥4 prior to TSE.

Finally, we also tested the effect of opioid administration in the ED (binary indicator) as an additional covariate in our fully adjusted model.

### Statistical Analyses

All statistical analyses were conducted in R (version 4.3.2, human cohort analyses) or GraphPad Prism (version 10.3.0, animal-based analyses). Prism files and code for all R-based analyses can be found at doi.org/10.15139/S3/XB2OJC.

Descriptive statistics for socio-demographic, ED/trauma-related, and past pain/stress variables were calculated for the full analysis cohort as well as for each CPMP trajectory group. To test for differences in socio-demographic characteristics between each CPMP trajectory, χ^2^ and Kruskal-Wallis χ^2^ comparisons were used. We tested the correlation of all ELA subscores using Spearman’s rank-order correlation. We also assessed for differences in reported rates of ELA by sex, race/ethnicity, ADI, and PDI using appropriate bivariate tests (e.g. Welch’s t-test, Kruskal-Wallis χ, or Spearman correlation).

To assess rates of ELA based on CPMP trajectory we first used bivariate Kruskal-Wallis tests. We then used multinomial logistic regression analyses to estimate the relative odds of following each latent CPMP trajectory based on composite CTQ score while controlling for standard demographic characteristics (age, sex, race, ED region). Then, we tested a more complete, adjusted model that accounted for additional demographic factors (age, sex, race, ED region, BMI, marital status, and ADI), trauma-related variables (trauma type, PDI), and past pain (pre-TSE moderate-severe pain) covariates. Though we also tested this fully adjusted model for the influence of opioid administration in the ED, we decided not to include this variable in subsequent analyses because there was a significant amount of missing data that further reduced the cohort sample size. Next, we calculated relative risk and population attributable fraction for ELA exposure and pain trajectory classification[49]. All independent variables included in the model were checked for multicollinearity using the Variance Inflation Factor (VIF). No selected variables had an adjusted generalized VIF>5, so all were retained for analysis. Interaction variables for CTQ × sex and CTQ × race/ethnicity were also tested in this fully adjusted model.

Each ELA subscore, including bullying, was tested individually in the fully adjusted regression model to identify the strongest subtypes of ELA. To compare across dissimilar subscore scales, we conducted additional analyses using binary indicator variables for a subscore≥1 as well as with z-score normalized subscore variables. Lastly, a top ELA predictor composite score was calculated by summing the top three predictors of pain trajectory. The fully adjusted model was run using this newly created predictor variable. All logistic regression analyses were conducted in reference to the low pain latent class. Benjamini-Hochberg (BH)-correction was applied both within and between models, as appropriate.

## Rodent Studies

### Animals

All animal studies were completed in accordance with the Institutional Animal Care and Use Committee (IACUC) at the University of North Carolina at Chapel Hill (UNC-CH) under an approved protocol (#21.256). Pregnant Sprague Dawley rat dams (gestational day 13) were purchased from Charles River Laboratories (Wilmington, MA, USA) and singled housed within the Division of Laboratory Animal Medicine at UNC-CH. The dams had *ad libitum* food and water and were on a 12-hour light/dark cycle (lights on at 07:00). Starting on gestational day 20, dams were monitored twice daily (08:00 and 18:00) for parturition. At parturition (notated as postnatal day 0; P0), pups were counted and sexed.

### Neonatal limited bedding model of early life adversity

We used the previously published and validated neonatal limited bedding (NLB) model of ELA[31; 32; 43; 82]. Each litter was randomly assigned to either -NLB (no NLB controls, n= 7 litters) or +NLB groups (n=7 litters). On P2, +NLB litters were transferred into a new cage with a wire grate elevated 2 cm above the cage bottom, devoid of standard wood-chip bedding material. These litters were given a single paper towel (25 x 25 cm) to use as bedding material. On P9, +NLB litters were returned to standard housing. -NLB control litters were moved into new cages with standard bedding on P2 and again on P9 to match the level of handling experienced by the +NLB litters. In addition to standard enrichment (acrylic rat tunnels and shreddable bags full of crinkle paper) -NLB litters were also given the same paper towel on P2 to control for the addition of a novel enrichment device. Following NLB, each litter was weaned on P21 and then reared in standard grouped housing (with their same sex, same group littermates) until further experiments at P120 where each animal was moved into single housing.

### Single Prolonged Stress

The SPS model has been used previously to model a single timepoint TSE[51; 53; 86], and was combined here with NLB to assess the effect of ELA on adult TSE and subsequent mechanical hypersensitivity. Briefly, at P120, animals from both the -NLB and +NLB groups were randomly assigned to a handled control group (-SPS) or an SPS-exposure group (+SPS), creating a two-by-two design with the following groups: -NLB/-SPS, +NLB/+SPS, -NLB/+SPS, and +NLB/-SPS. SPS was performed on male and female animals as previously described[86], which included exposure to three different stressors within one session: 2 hours of restraint, 20 minutes of forced swim (followed by 20 minutes of recovery), and exposure to diethyl ether until sedation was achieved. They were then monitored until the return of their righting response. Following handling or SPS-exposure, all animals were returned to the housing facility and were single housed from this point forward.

### Von Frey Testing

All behavior experiments were conducted by a single experimenter, under light conditions Animals were acclimated to the testing apparatus over a period of four days in the week prior to behavioral assessments. On these days, each rat was placed in a testing chamber for 30 minutes and a small blunt probe was applied to the paw, such that a withdrawal response was not elicited. To estimate mechanical hypersensitivity (an approximation of pain-like behavior) in our rodents, we used an electronic von Frey system (WPI, Sarasota, FL, USA). Prior to the start of each von Frey session, animals were acclimated to their testing environment for 30 minutes. Then, von Frey measurements (paw withdrawal threshold; PWT) were taken starting at 09:00 by applying a blunt von Frey filament probe to the right hind paw with accumulating force until the animal withdrew its paw. This was performed five times with a five-minute rest period between each reading. Pre-SPS PWTs were taken on two days prior to SPS. Starting 24 hours after SPS, PWTs were measured for nine days and then again on days 12, 14, 16, and 20 post-SPS. SPS-unexposed rats were tested on the same days as their SPS-exposed counterparts. The behavioral scientist for these experiments was blinded to which animals were exposed to NLB.

### Statistical Analysis

PWT replicates were processed to remove outliers and calculate daily average PWT. To test for group differences in PWT prior to SPS, we conducted a Welch-corrected t-test. To test for the effect of time following SPS and its interaction with NLB, we used a mixed model analysis for repeated measures. Following identification of a significant interaction, we conducted post hoc Tukey’s multiple comparison tests at each timepoint. Statistical analyses were conducted using Graphpad Prism (version 10.3.0). Prism files associated with these analyses are deposited at doi.org/10.15139/S3/XB2OJC.

## RESULTS

### Study design

Translational studies from both humans and animals were used in the current set of analyses (**Figure 1**). In humans, study participants were enrolled in EDs from across the US within 72 hours of a TSE.[62]. ELA was retrospectively reported in a clinical interview session two weeks following discharge from the ED. Pain severity was also self-reported at regular intervals following ED discharge, and these data were used to develop pain trajectory groups, as previously described.[6; 72] In animals, we combined the SPS model, which has previously been shown to cause enduring mechanical hypersensitivity[36; 52; 77; 86; 93], with the NLB model of ELA. We then tested the effect of NLB and SPS on mechanical hypersensitivity over time.

**Figure 1:**
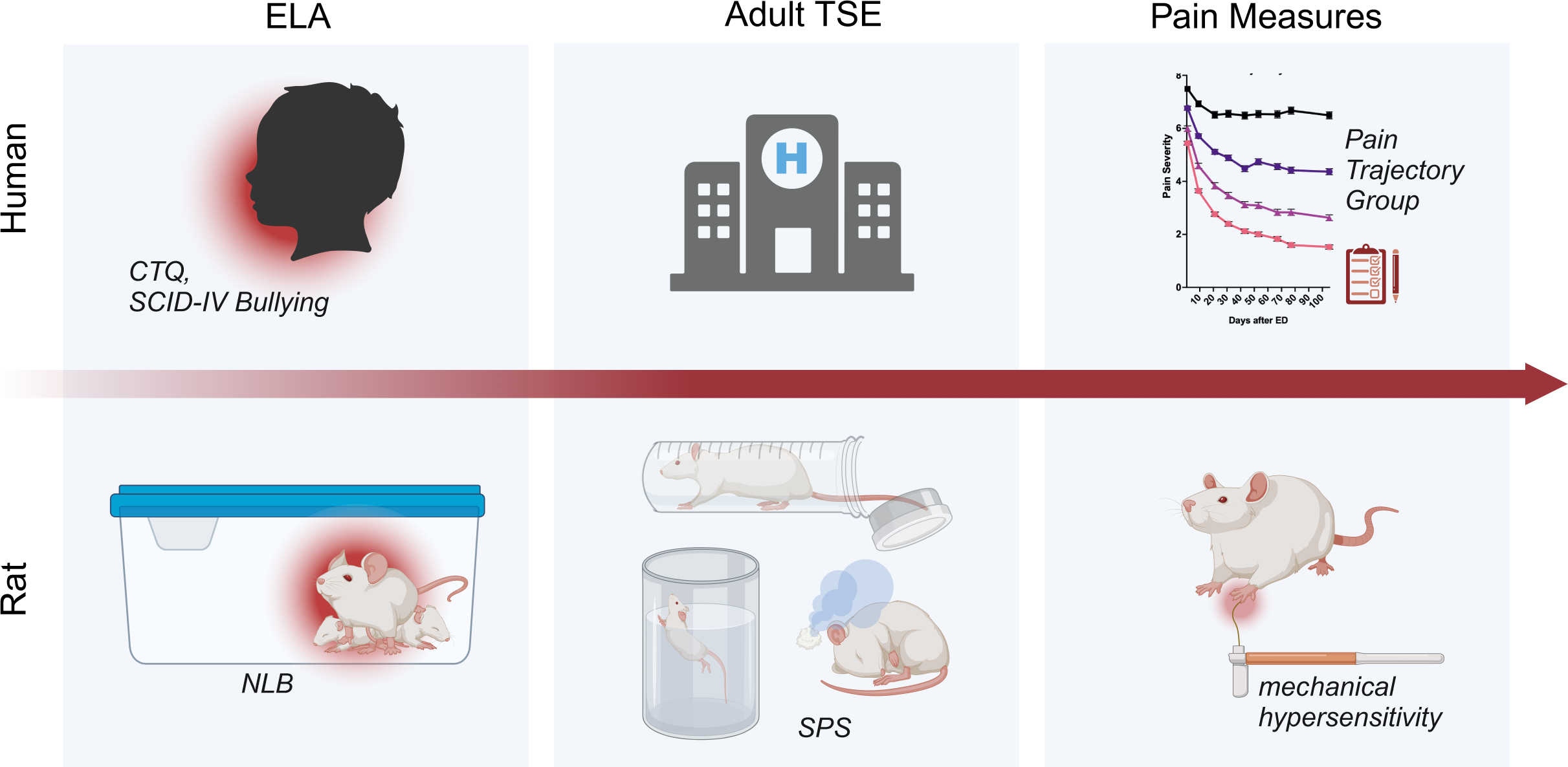
This translational research study used data from a prospective human cohort study of TSE survivors along with parallel animal experiments to test the effect of ELA on pain measures following adult TSE. (Top) The AURORA study recruited participants who reported to an ED within 72 hours of TSE. As part of the AURORA study, participants provided retrospective data about past ELA (CTQ and bullying) via standard questionnaires. Analysis of participant pain outcome data identified four pain trajectories spanning the eight weeks following TSE and these trajectories were used to test the association between ELA and pain following TSE. (Bottom) In animals, Sprague Dawley rat pups were exposed to -NLB or +NLB from PD2-9. In adulthood, rats were exposed to the TSE model, SPS, and pain-related mechanical hypersensitivity was assessed using electronic von Frey. *TSE-traumatic stress event*, *ELA-early life adversity*, *ED-emergency department*, *CTQ-childhood trauma questionnaire*, *NLB-neonatal limited bedding*, *PD-postnatal day*, *SPS-single prolonged stress*. *Created in BioRender. Verma, T. (2024) BioRender.com/o59q848*

### Sample characteristics

Baseline participant characteristics are shown in **Table 1**. A total of n=2,480 men and women from the AURORA study had available study data and were included in the current analyses. Most participants were women (n=1,554, 63%), non-Hispanic Black (n=1,213, 49%), and experienced MVC-related trauma (n=1,909, 77%). The average age was 36.1±13.3 and the average BMI was 30.1±8.3. The population was largely disadvantaged (ADI=63.9±27.6) and a minority of participants had moderate pain in the 30 days prior to TSE (NRS≥4, n=777, 31%).

**Table 1.** Participant characteristics.

|  | Total Sample (N = 2,480) |  |
| --- | --- | --- |
|  | % or Mean | (n) or (SD) |
| **Socio-Demographic Variables** |  |  |
| Female | 63% | 1,554 |
| Age | 36.09 | 13.31 |
| Race/Ethnicity |  |  |
| Hispanic | 11% | 278 |
| Non-Hispanic white | 36% | 891 |
| Non-Hispanic Black | 49% | 1,213 |
| Other | 4% | 91 |
| BMI | 30.13 | 8.32 |
| Area Deprivation Index | 63.93 | 27.57 |
| **ED/Trauma-Related Variables** |  |  |
| Site |  |  |
| Northeast | 42% | 1,053 |
| Southeast | 19% | 467 |
| Midwest | 39% | 960 |
| Trauma Type |  |  |
| MVC | 77% | 1,909 |
| Physical/Sexual Assault | 9% | 231 |
| Other | 14% | 340 |
| Peritraumatic Distress Inventory | 13.92 | 7.24 |
| **Past Pain/Stress Variables** |  |  |
| Had Previous Moderate/Severe Pain | 31% | 777 |
| Childhood Trauma Questionnaire (CTQ) |  |  |
| Any Childhood Trauma (CTQ≥1) | 80% | 1,977 |
| CTQ Composite Score (0-44) | 9.50 | 9.79 |
| Any Physical Abuse | 44% | 1,089 |
| Physical Abuse Subscore (0-8) | 1.56 | 2.31 |
| Any Emotional Abuse | 66% | 1,627 |
| Emotional Abuse Subscore (0-8) | 2.56 | 2.64 |
| Any Sexual Abuse | 35% | 867 |
| Sexual Abuse Subscore (0-12) | 1.93 | 3.35 |
| Any Physical Neglect | 46% | 1,146 |
| Physical Neglect Subscore (0-8) | 1.53 | 2.14 |
| Any Emotional Neglect | 52% | 1,284 |
| Emotional Neglect Subscore (0-8) | 1.92 | 2.36 |
| Bullying |  |  |
| Any | 80% | 1,977 |
| Bullying Score (0-8) | 2.97 | 2.37 |

*SD*-standard deviation; *BMI*-body mass index; *ED*-emergency department; *MVC*-motor vehicle collision; *CTQ*-childhood trauma questionnaire; *ELA*-early life adversity.

### ELA rates in the AURORA study

Most participants (n=2,260, 91%) reported experiencing at least one instance of any type of ELA (i.e., either CTQ≥1 or childhood Bullying ≥1). The most common CTQ subtype was emotional abuse (n=1,627, 66%), followed by emotional neglect (n=1,284, 52%). However, the largest proportion of individuals reporting past ELA experienced bullying in childhood (n=1,977, 80%). In addition, most participants (n=1,906, 77%) reported experiencing more than one subtype of ELA, and all subtypes of ELA were statistically significantly correlated with each other (*p*<0.001, ***Supplementary Figure 1***). The most highly correlated pairs of ELA subtypes were physical and emotional neglect (Spearman’s ρ=0.83, *p*<0.001) and physical and emotional abuse (Spearman’s ρ=0.72, *p*<0.001). All other pairs of ELA subtypes had a Spearman’s ρ value between 0.18 and 0.59, *p*<0.001.

Because of previous studies indicating sex or gender differences in the experience of ELA[20; 33], we assessed ELA rates in women versus men in the AURORA cohort. Women reported higher levels of ELA than men when examining both CTQ composite score (Welch’s t (2,265.3)=5.97, *p*<0.001, women: 10.4±10.4, men: 8.1±8.4) and physical, emotional, and sexual abuse ELA subtypes (***Supplementary Figure 2***). However, men reported higher levels of physical neglect (Welch’s t(1,876.9)=-2.03, *p*=0.042), and both sexes reported similar levels of emotional neglect and bullying (*p*>0.05). In addition to sex differences, we also assessed whether CTQ scores differed by race/ethnicity groups, as has been reported previously[58], but we only found a trend-level statistically significant relationship (Kruskal-Wallis χ (3)=7.79, *p*=0.051, ***Supplementary Figure 3)*** indicating that Hispanic individuals had marginally higher rates of ELA compared to non-hispanic white, non-hispanic black, and “other” groups. Positive, but modest, correlations were identified between both ADI and CTQ (Spearman’s ρ=0.08, *p*<0.001) and PDI and CTQ (Spearman’s ρ=0.17, *p*<0.001).

### CPMP prevalence in the AURORA study as assessed via latent class trajectory groups

CPMP development versus recovery was defined as described previously[6; 72] by four latent classes including a low pain class (n=1,070, 43%), a moderate recovery pain class (n=468, 19%) a moderate unremitting pain class (n=590, 24%), and a high unremitting pain class (n=352, 14%) (**Figure 2A**). Participant characteristics corresponding to each post-TSE pain trajectory group are shown in ***Supplementary Table 1***. Average post-TSE pain NRS scores over time for each of the CPMP trajectories are shown in **Figure 2A**.

**Figure 2.**
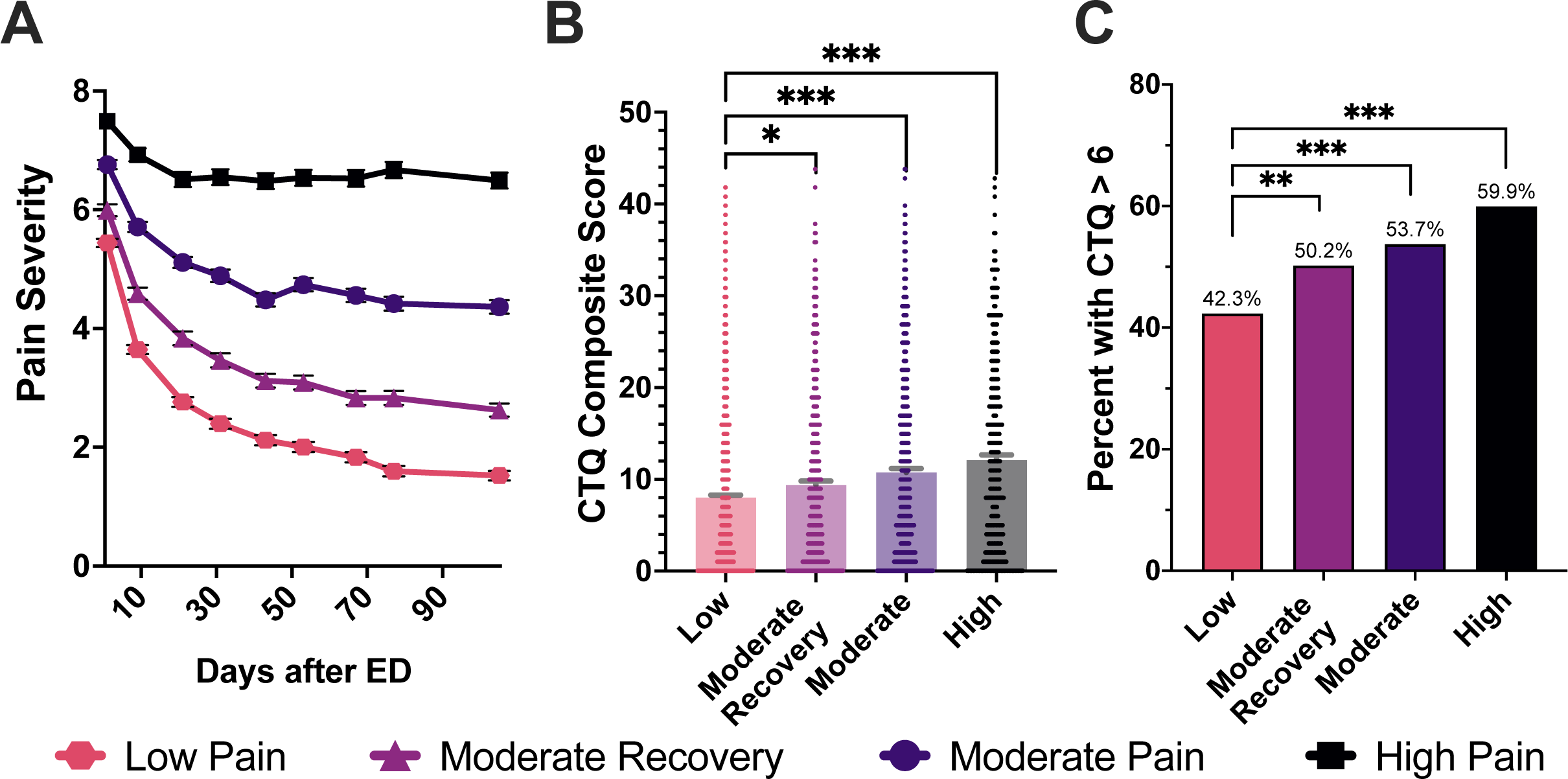
CPMP latent class trajectory groups and the relationship between the four latent class groups and CTQ. **A**) The four latent class trajectory groups in AURORA study participants including a high pain, moderate pain, moderate recovery, and low pain class with average pain scores graphed over a 3 month period following traumatic stress exposure (0-10 NRS, +/-SEM). **B)** Average CTQ composite scores in each latent class, **C)** percentage of individuals with a CTQ score above the median (CTQ>6) in each latent class group are shown. Bars show mean and error bars show standard error of the mean. Individual values are represented by dots overlaid onto the bar charts. *Abbreviations: NRS-Numerical Rating Scale, ED-emergency department, CTQ-childhood trauma questionnaire, CPMP-chronic posttraumatic musculoskeletal pain. Significance:*p<0.05, **p<0.01, ***p<0.001, ****p<0.0001*

### Higher amounts of ELA were associated with increased CPMP

Before evaluating the relationship between ELA and chronic pain trajectories following TSE, we first tested whether participants’ retrospective reports of pain before their TSE depended on their past ELA severity. We found that there was a statistically significantly increased odds of having previous moderate-to-severe pain with both higher CTQ composite (OR=1.032, p<0.001) and bullying scores (OR=1.096, p<0.001). We also conducted these analyses using z-score normalized predictor variables, enabling comparisons across predictors. The odds ratio for the z-score normalized CTQ composite score and bullying scores were 1.368 and 1.245, respectively (*p*<0.001).

In our primary study analyses, we first assessed the relationship between overall CTQ scores and post-TSE pain trajectories using bivariate analyses. We found a statistically significant difference in overall CTQ scores between the four CPMP trajectory groups (Kruskal-Wallis χ (3)=57.70, *p*<0.001, **Figure 2B**) with a mean CTQ score of 12.1±10.6 for high pain, 10.8±10.3 for moderate pain, 9.4±9.5 for moderate recovery, and 8.0±9.0 for low pain trajectories. In addition, a larger percentage of individuals with substantial CTQ (i.e. CTQ>6, the median CTQ score in the population) belonged to the high pain class (n=211, 60%; χ (1)=32.99, *p*<0.001), the moderate pain class (n=317, 54%; χ^2^(1)=19.85, *p*<0.001), or the moderate recovery pain class (n=235, 50%; χ (1)=8.17, *p*=0.004), than the low pain class (n=453, 42%; **Figure 2C**).

We found similar trends for childhood bullying across the CPMP trajectories (Kruskal-Wallis χ (3)=52.79, *p*<0.001, ***Supplementary Figure 4 A*** and ***B***. We also found that experiencing any ELA increased the relative risk for being in the high pain trajectory by 68.6% compared to the low pain trajectory group (RR=1.686, *p*<0.001). In addition, population attributable fraction suggested that we could expect a 34.8% reduction in high pain trajectory members if ELA exposure in the population was reduced to zero (***Supplementary Table 2***).

We then used multinomial logistic regression analyses to assess the relationship between CTQ and pain class membership while controlling for age, sex, race/ethnicity, and ED region and found that for every additional point on the composite CTQ score (range 0-44), an individual had 4.7% increased odds of being in the high pain trajectory group compared to the low pain trajectory group (OR=1.047, *p*<0.001, Benjamini-Hochberg (BH)-corrected *p*<0.001) (**Table 2**). The odds of being in the moderate or moderate recovery groups were also increased compared to the low pain group but to incrementally lesser amounts (moderate pain OR=1.031, *p*<0.001, BH-corrected *p*<0.001; moderate recovery OR=1.018, *p*=0.004, BH-corrected *p*=0.011).

**Table 2.** Model Summary for ELA Predictor Variables.

| *\*low pain group as reference* | **Moderate Recovery** |  |  | **Moderate Pain** |  |  | **High Pain** |  |  |
| --- | --- | --- | --- | --- | --- | --- | --- | --- | --- |
|  | OR | (95% CI) | *BH-adj. p* | OR | (95% CI) | *BH-adj. p* | OR | (95% CI) | *BH-adj. p* |
| Model 1 (CTQ Composite) | 1.018 | 1.006 - 1.030 | **0.011** | 1.031 | 1.020 - 1.042 | **<0.001** | 1.047 | 1.034 - 1.060 | **<0.001** |
| *Adjusts for age, site, race, gender on birth certificate,* |  |  |  |  |  |  |  |  |  |
| Model 2 (CTQ Composite, full covariates) | 1.013 | 1.000 - 1.026 | 0.119 | 1.022 | 1.010 - 1.035 | **0.001** | 1.030 | 1.016 - 1.045 | **<0.001** |
| *Adjusts for Model 1 adjusters and BMI, previous moderate/severe pain, marital status, Area Deprivation Index, trauma type, and Peritraumatic Distress Inventory* |  |  |  |  |  |  |  |  |  |
| Model 3 (top 3 ELA predictors, full covariates) | 1.021 | 1.000 - 1.042 | 0.120 | 1.045 | 1.025 - 1.065 | **<0.001** | 1.068 | 1.044 - 1.092 | **<0.001** |
| *Used new composite score for only physical abuse, emotional abuse, and bullying* |  |  |  |  |  |  |  |  |  |

CTQ-childhood trauma questionnaire score; OR-odds ratio; CI-confidence interval; BH-Benjamini Hochberg; BMI-body mass index
Significant *p* -values are shown in bold

When we controlled for additional demographic, pre-trauma, and trauma-related covariates, including marital status, BMI, previous moderate-to-severe pain, ADI, trauma type, and PDI, CTQ was significantly associated with increased odds of being in each of the three CPMP classes, though the moderate recovery group did not survive multiple comparisons correction: high pain (OR=1.030, *p*<0.001, BH-corrected *p*<0.001), moderate (OR=1.022, *p*<0.001, BH-corrected *p*=0.001), moderate recovery (OR=1.013, *p*=0.047, BH-corrected *p*=0.119)(**Table 2**). In this full model, we also tested opioid administration in the ED as a covariate because of its potential influence on both pain and posttraumatic stress outcomes.[55; 63] Opioid use did not substantially influence effect size estimates or statistical significance for the relationship between CTQ and CPMP in either the severe or moderate pain classes (*p*>0.05; ***Supplementary Table 3***). Because opioid use in the ED reduced our sample size substantially, all subsequent analyses did not include this measure.

Because we found differences in the level of CTQ in men and women, in secondary analyses we assessed for an interaction in our complete model between CTQ and sex but did not find a significant interaction (*p>*0.05). Similarly, we tested for an interaction between race/ethnicity and CTQ on CPMP trajectories using our full covariate model and did not find a significant interaction (*p*>0.05).

### Physical and emotional abuse and childhood bullying were the strongest predictors of post-TSE CPMP trajectories

Using the fully adjusted model described above, we independently tested each CTQ subtype and bullying score to identify which type of ELA was most significantly associated with CPMP. We first conducted these analyses using each subscale score as a predictor, which enabled interpretation of odds ratios in relation to a single point increase in that ELA subscore. The CTQ abuse subtypes (physical, emotional, and sexual) and bullying were associated with significantly increased odds of being in the moderate pain or high unremitting pain class (*p*<0.05), but the effect for sexual abuse did not survive BH correction (**Table 3**). Physical and emotional neglect subtypes were not associated with any pain latent class trajectory (*p*>0.05). The highest odds for being in the high unremitting pain class was observed for emotional abuse (OR=1.167, *p*<0.001, BH-corrected *p*<0.001), followed closely by bullying (OR=1.161, *p*<0.001, BH-corrected *p*<0.001), and physical abuse (OR=1.151, *p*<0.001, BH-corrected *p*<0.001).

**Table 3.** ELA Subscores Model Summary in Full Covariate Model.

| * low pain as reference group<br>ELA Subtype | Moderate Recovery |  |  | Moderate Pain |  |  | High Pain |  |  |
| --- | --- | --- | --- | --- | --- | --- | --- | --- | --- |
|  | OR | 95% CI | BH-adj. p | OR | 95% CI | BH-adj. p | OR | 95% CI | BH-adj. p |
| Physical abuse |  |  |  |  |  |  |  |  |  |
| CTQ subscore | 1.012 | 0.956 - 1.071 | 0.980 | 1.102 | 1.049 - 1.159 | **0.003** | 1.151 | 1.085 - 1.220 | **<0.001** |
| any (subscore≥1) | 1.187 | 0.927 - 1.519 | 0.682 | 1.613 | 1.275 - 2.041 | **0.002** | 1.758 | 1.312 - 2.357 | **0.004** |
| Sexual abuse |  |  |  |  |  |  |  |  |  |
| CTQ subscore | 1.043 | 1.004 - 1.085 | 0.226 | 1.057 | 1.020 - 1.095 | **0.046** | 1.057 | 1.013 - 1.102 | 0.114 |
| any (subscore≥1) | 1.474 | 1.140 - 1.906 | 0.061 | 1.407 | 1.100 - 1.799 | 0.075 | 1.356 | 1.001 - 1.837 | 0.333 |
| Emotional abuse |  |  |  |  |  |  |  |  |  |
| CTQ subscore | 1.056 | 1.006 - 1.108 | 0.219 | 1.110 | 1.061 - 1.162 | **<0.001** | 1.167 | 1.104 - 1.233 | **<0.001** |
| any (subscore≥1) | 1.510 | 1.164 - 1.958 | **0.036** | 1.546 | 1.204 - 1.985 | **0.014** | 1.753 | 1.273 - 2.413 | **0.014** |
| Physical neglect |  |  |  |  |  |  |  |  |  |
| CTQ subscore | 1.013 | 0.957 - 1.073 | 0.980 | 1.028 | 0.974 - 1.084 | 0.805 | 1.047 | 0.980 - 1.118 | 0.676 |
| any (subscore≥1) | 1.191 | 0.932 - 1.521 | 0.629 | 1.323 | 1.046 - 1.674 | 0.171 | 1.355 | 1.010 - 1.818 | 0.311 |
| Emotional neglect |  |  |  |  |  |  |  |  |  |
| CTQ subscore | 1.051 | 1.000 - 1.106 | 0.385 | 1.022 | 0.973 - 1.073 | 0.805 | 1.044 | 0.983 - 1.108 | 0.643 |
| any (subscore≥1) | 1.220 | 0.958 - 1.555 | 0.498 | 1.247 | 0.987 - 1.575 | 0.445 | 1.261 | 0.941 - 1.689 | 0.512 |
| Bullying |  |  |  |  |  |  |  |  |  |
| Subscore | 1.065 | 1.010 - 1.123 | 0.169 | 1.093 | 1.039 - 1.149 | **0.011** | 1.161 | 1.092 - 1.234 | **<0.001** |
| any (subscore≥1) | 1.611 | 1.164 - 2.231 | 0.074 | 1.164 | 0.873 - 1.551 | 0.785 | 1.247 | 0.866 - 1.796 | 0.785 |

ELA-early life adversity; OR-odds ratio; CI-confidence interval; BH-Benjamini Hochberg; CTQ-childhood trauma questionnaire score
Significant *p* -values are shown in bold

We then tested whether a binary indicator variable for any amount of each ELA subtype (i.e. subscore≥1) was a predictor of CPMP trajectory. We found that having any history of childhood physical abuse was associated with 75.8% increased odds of experiencing high unremitting CPMP (*p*<0.001, BH-corrected *p*=0.004) and 61.3% increased odds of experiencing moderate CPMP (*p*<0.001, BH-corrected *p*=0.002). Similar to physical abuse, experiencing any early life emotional abuse was associated with 75.3% increased odds of experiencing high unremitting CPMP (*p*<0.001, BH-corrected *p*=0.014) and 54.6% increased odds for moderate CPMP (*p*<0.001, BH-corrected *p*=0.014). The binary predictor variable for any past bullying, however, was not a significant predictor of any pain class following BH-correction (BH-corrected *p*>0.05). Finally, because some of the six ELA subscores had different score ranges, we ran our fully adjusted model with z-score normalized subscores enabling direct comparison of each type of ELA (***Supplementary Table 4***). Compared to the other ELA subscores, emotional abuse (high CPMP: OR=1.502, *p*<0.001, BH-corrected *p*<0.001; moderate CPMP: OR=1.317, *p*<0.001, BH-corrected *p*<0.001) and bullying (high CPMP: OR=1.425, *p*<0.001, BH-corrected *p*<0.001; moderate CPMP: OR=1.234, *p*=0.001, BH-corrected *p*=0.011) were the two strongest predictors of high and moderate CPMP development, but not of moderate recovery (emotional abuse: OR=1.154, *p*=0.028, BH-corrected *p*=0.230; bullying: OR=1.161, *p*=0.020, BH-corrected *p*=0.191). Physical abuse followed closely with OR=1.382 (*p*<0.001, BH-corrected *p*<0.001) for high CPMP and OR=1.251 (*p*<0.001, BH-corrected *p*=0.003) for moderate CPMP. Using z-score normalized subscores, sexual abuse, physical neglect, and emotional neglect were not significantly associated with CPMP trajectory (*p*>0.05).

Building upon the finding that physical abuse, emotional abuse, and bullying were the strongest predictors of CPMP, we tested our full model using a newly computed composite score consisting of only these three variables (**Table 2**). The odds ratio for this new composite score was 1.068 in the high pain group (compared to the low pain group, *p*<0.001, BH-corrected *p*<0.001), 1.045 in the moderate pain group (*p*<0.001, BH-corrected *p*<0.001), and 1.021 in the moderate recovery group (*p*=0.051, BH-corrected *p*=0.120).

### In animals, ELA increases the severity of baseline mechanical hypersensitivity

In male and female rats, we found that +NLB rats without SPS exposure exhibited statistically significant decreased PWT compared to -NLB rats without SPS exposure (n=42, M±SEM=35.16±0.80 versus M±SEM=41.71±1.72; Welch’s corrected t(24.20)=3.44, *p*=0.006; **Figure 3A**).

**Figure 3.**
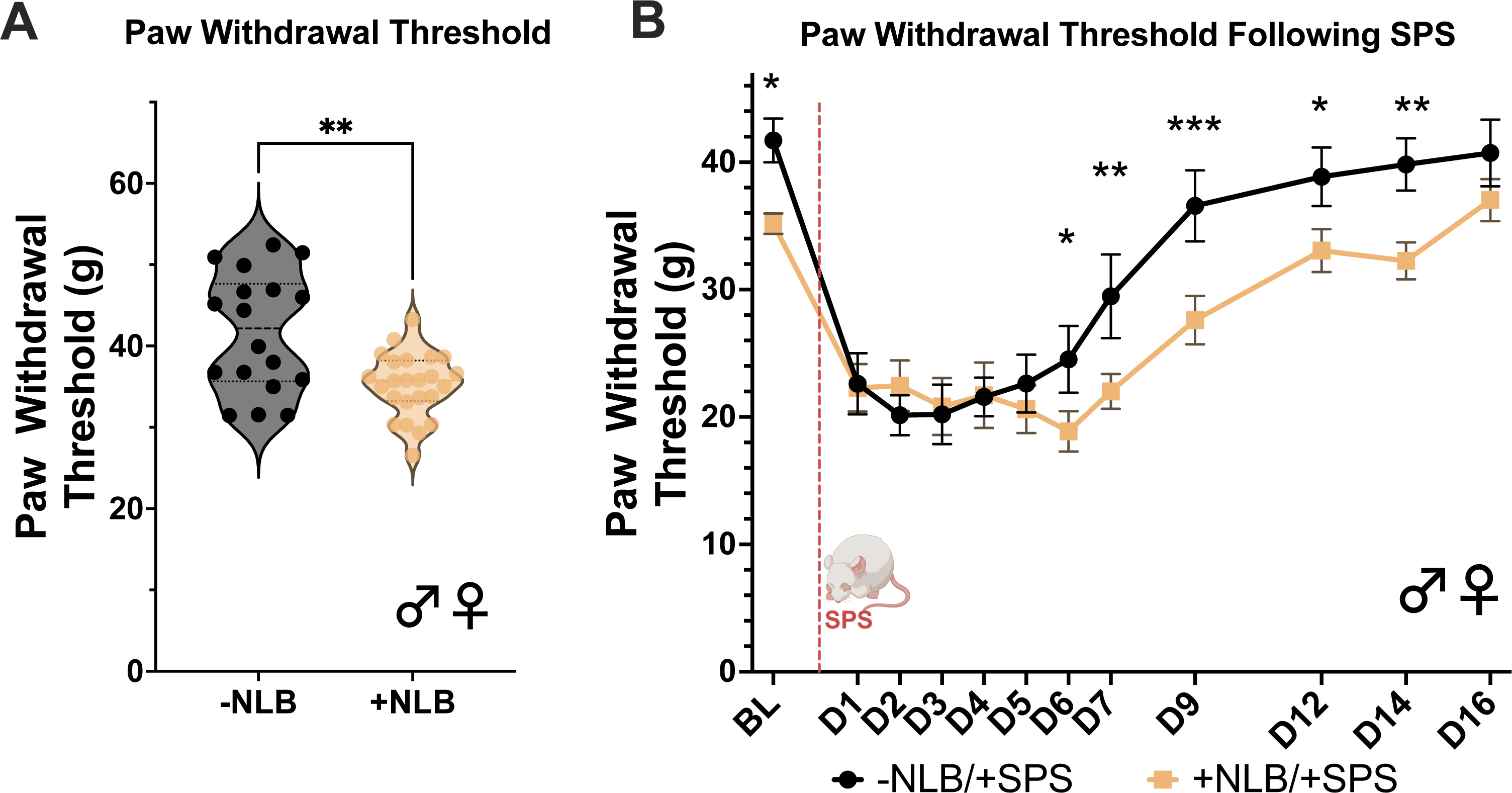
Paw withdrawal threshold in the NLB and SPS rat models of ELA and TSE **A**) +NLB significantly decreases paw withdrawal threshold in male and female rats (N=42) compared to - NLB. **B)** Single prolonged stress decreases paw withdrawal threshold in both -NLB (n=18) and +NLB (n=24) rats, but -NLB rats return to baseline paw withdrawal threshold earlier than +NLB. From D6-14 following SPS, +NLB and -NLB rats’ paw withdrawal thresholds were significantly different from days 6-14. Dots and squares represent means and error bars represent standard error of the mean. *Abbreviations: NLB-neonatal limited bedding, SPS-Single Prolonged Stress, g-grams, ELA-early life adversity, TSE-traumatic stress exposure, D6-day 6 following SPS. Significance:*p<0.05, **p<0.01, ***p<0.001*

### ELA extends the duration of mechanical hypersensitivity following adult TSE

Following baseline mechanical hypersensitivity assessments, we assessed the duration of enduring mechanical hypersensitivity following adult SPS in +NLB versus -NLB animals. We found that male and female +NLB rats exhibited a longer duration of mechanical hypersensitivity compared to -NLB animals (**Figure 3B**). Specifically, we found a significant interaction between NLB and time post-SPS when assessing PWT (F(11,197)=3.220, *p*<0.001). Tukey’s multiple comparison test revealed that +NLB/+SPS rats had significantly lower PWTs than -NLB/+SPS rats on days 6 (*p-adj.*=0.022), 7 (*p-adj.*=0.003), 9 (*p-adj.*<0.001), 12 (*p-adj.*=0.012), and 14 (*p-adj.*=0.002) following SPS (**Figure 3B**).

## DISCUSSION

Over the past few decades, ELA has been shown to increase adult vulnerability to many adverse health outcomes including substance use disorder[1; 23], heart disease[4; 68], and sleep disorders[44], among others. Here, we add to this literature by demonstrating in both a large longitudinal human cohort study and a back-translated animal model, that ELA also prolongs the duration of chronic pain following adult TSE suggesting increased latent vulnerability to CPMP in individuals with ELA. In human TSE survivors, we demonstrated this relationship using overall CTQ scores, childhood bullying scores, and latent class trajectory groups representing the four main pain outcome groups in AURORA study participants. We also assessed each subtype of CTQ (and bullying) and determined that physical abuse, emotional abuse, and bullying were most associated with CPMP duration. Finally, we showed that observations from our human cohort study could be modeled effectively in rodents by repurposing a combination of well-studied ELA and TSE models. Development of such an animal model is highly valuable for examining mechanisms by which ELA creates vulnerability to CPMP after traumatic stress exposures during adulthood.

Results from our human data analyses support our hypothesis that ELA is associated with increased risk for CPMP and are consistent with previous literature showing a dose-response effect of increased ELA on increased pain in adulthood.[15; 16; 37; 91] We also extended the field’s general understanding of additional factors related to TSE survivors’ experience with ELA. For example, we showed that the overall rates of ELA are higher than the US national average[30; 40] in adult trauma survivors who report for care in the immediate aftermath of TSE. These data are consistent with previous publications documenting increased ELA in the broader emergency department population.[12; 22; 57; 73] Additionally, we found that most individuals in this population experience at least some bullying in adolescence and, consistent with previous reports, women report the highest rates of ELA[30; 33]. Our results indicating that physical abuse is one of the strongest predictors of CPMP is consistent with the results of a recent large meta-analysis of 57 studies showing that physical abuse is the ELA subtype most associated with chronic pain in adulthood[16] and is interesting in comparison to a previous analysis of the AURORA study showing that other types of ELA (compared to physical abuse) are more strongly associated with neuropsychiatric outcomes of TSE[89; 94] such as posttraumatic stress disorder and major depressive disorder[94].

Similar to our AURORA-based finding showing the highest CTQ scores in participants who did not experience pain improvement by 6 months post-TSE, the NLB+SPS model showed a longer duration of mechanical hypersensitivity compared to SPS alone. The NLB model captures the ‘emotional’/stress aspects of ELA[82] but does not consider the effects of tissue injury in childhood that occur in instances of early life physical abuse. Because our human studies indicated that physical abuse is a top predictor of CPMP (but not a top predictor of posttraumatic stress and depressive symptoms[94]), future experiments should assess whether the addition of physical injury at the time of NLB lengthens the duration or increases the severity of mechanical sensitivity in male and female rats after adult TSE. A few studies have shown that the addition of tissue injury to SPS in adulthood, such as plantar incision or spared nerve injury, leads to increased mechanical sensitivity[8] and others have shown that neonatal tissue injury alone increases pain outcomes in preclinical studies[83; 88], but there is a lack of literature showing the combined effects of NLB+neonatal injury on adult hypersensitivity after TSE. Such models could help improve the face validity of animal ELA experiments in modeling its relationship with CPMP.

While the current study did not assess mechanisms through which ELA increases risk for CPMP, multiple studies assessing similar relationships have addressed this topic previously.[5; 18; 34; 54] Therefore, we can draw from these other studies, which implicate enduring ELA-induced changes to immune and neuroendocrine stress interactions[5; 65], to develop hypotheses for future studies assessing the relationship between ELA and CPMP. For example, a previous meta-analysis showed the inflammatory marker, CRP, and pro-inflammatory cytokines, TNF-α and IL-6, are elevated in individuals with ELA across 25 studies.[5] Another study showed that cortisol levels were blunted after adult trauma in individuals with prior substantial stressors.[18] Finally, a third study showed that the relationship between ELA and chronic multisite pain was mediated by hippocampal size and genetic risk in the stress system gene *Fkbp5*[54]. This study is consistent with substantial previous research in the field of psychiatry showing the influence of ELA on *Fkbp5*/FKBP51 biology.[46; 79; 85] The differences in chronic pain observed in these studies and many others might be mediated at the level of the DNA by epigenetic changes (e.g. CpG methylation) to key genes (e.g. *Fkbp5*, *Slc6a4*)[38; 46] involved in downstream neuro-stress-immune processes that later confer the effects of an individual’s historical environment to their present-day biology and behavior.[34; 38]

These findings highlight the need to refine our understanding of the molecular mechanisms driving CPMP more generally. This is particularly important for individuals who have endured ELA, as they are the most highly burdened individuals, both in terms of expected adverse adult health outcomes and the other societal and environmental disadvantages they experience (e.g. racial/ethnic discrimination[58], adult socioeconomic deprivation[35], or natural disaster exposure[71]). Indeed, the body of literature showing a dose-dependent relationship between ELA and chronic pain[15; 16; 37; 91] supports the idea that higher counts of ELA confer a high degree of risk for later health outcomes. Similar patterns have also been noted in the neuropsychiatric literature.[42]

Strengths of this study include the use of translational data across both a large multiethnic human cohort study of TSE survivors and a parallel animal model experiment. Several limitations should also be considered when interpreting this study data. First, the types of ELA assessed in AURORA were limited to those presented here. Therefore, we do not know how additional or different stressors such as e.g. food insecurities, socioeconomic disadvantage, or loss of a parent or loved one would influence CPMP. Second, we do not know the timing of ELA experienced by AURORA participants. Multiple studies suggest that the timing of ELA in childhood can have a strong impact on adverse outcome development in adulthood[56; 74; 84; 92], however there is still a lack of consensus as to how developmental timing of ELA relates to specific outcomes, their severity, and their interplay with other factors such as sex or gender. Several studies support that early childhood ELA, in line with the timing of our animal model, may confer a particularly strong vulnerability for adverse adult health outcomes.[56; 74; 84] Nonetheless, as the field of ELA research continues to grow, developmental timing of ELA should be a high priority of future human and animal model studies. Third, in our animal studies we found that ELA only increased the duration and not the magnitude of mechanical sensitivity following SPS, while in humans ELA influenced both measures. Future studies should explore additional factors (e.g. tissue injury at the time of NLB and/or SPS) that could further improve the face validity of our animal model. Fourth, self-report measures of ELA are known to be imperfect.[67] However, they are the most practical tool to assess retrospective ELA history[60]. Finally, in our animal studies, we used only a reflexive measure of pain-like behavior, which does not fully model the entire human pain experience. Future studies would benefit from additional measures of mechanical hypersensitivity such as naturalistic or operant behaviors.[69]

In conclusion, we showed that ELA increases pain duration following adult TSE in both humans and animals. This work provides a substantial foundation on which to conduct future mechanistic studies assessing mechanisms that drive non-recovery in ELA survivors. Through such mechanistic discovery efforts, subsequent validation efforts, and further improvement of animal models of the ELA/CPMP relationship, we are hopeful that we and others can develop novel therapeutic strategies that target the unique pathobiology inherent to individuals with an adverse childhood, with the ultimate goal of reducing disparities in recovery processes following TSE.

## Data Availability

All data produced are available online at https://nda.nih.gov/edit_collection.html?id=2526.

## Acknowledgements

The investigators wish to thank the trauma survivors participating in the AURORA Study. Their time and effort during a challenging period of their lives make our efforts to improve recovery for future trauma survivors possible. This work was funded by the National Institute of Mental Health (NIMH; U01MH110925-PIs McLean, Kessler, Ressler, and Koenen), by the National Institute of Neurological Disorders and Stroke (NINDS; R01NS118563-PIs Linnstaedt and McLean), by The Rita Allen Foundation (PI Linnstaedt), by the National Institute of General Medical Sciences (NIGMS; T32GM008450-Fellow McKibben), as well as the US Army MRMC, One Mind, and The Mayday Fund. The content is solely the responsibility of the authors and does not necessarily represent the official views of any of the funders. Data used in this manuscript is available through the National Institute of Mental Health (NIMH) Data Archive (NDA). The NDA Collection for the AURORA Project can be found here: https://nda.nih.gov/edit_collection.html?id=2526. This manuscript reflects the views of the authors and may not reflect the opinions or views of the NIH or of the Submitters submitting original data to NDA.

