## Supplementary File for "Early life adversity increases risk for chronic posttraumatic pain, data from humans and rodents"

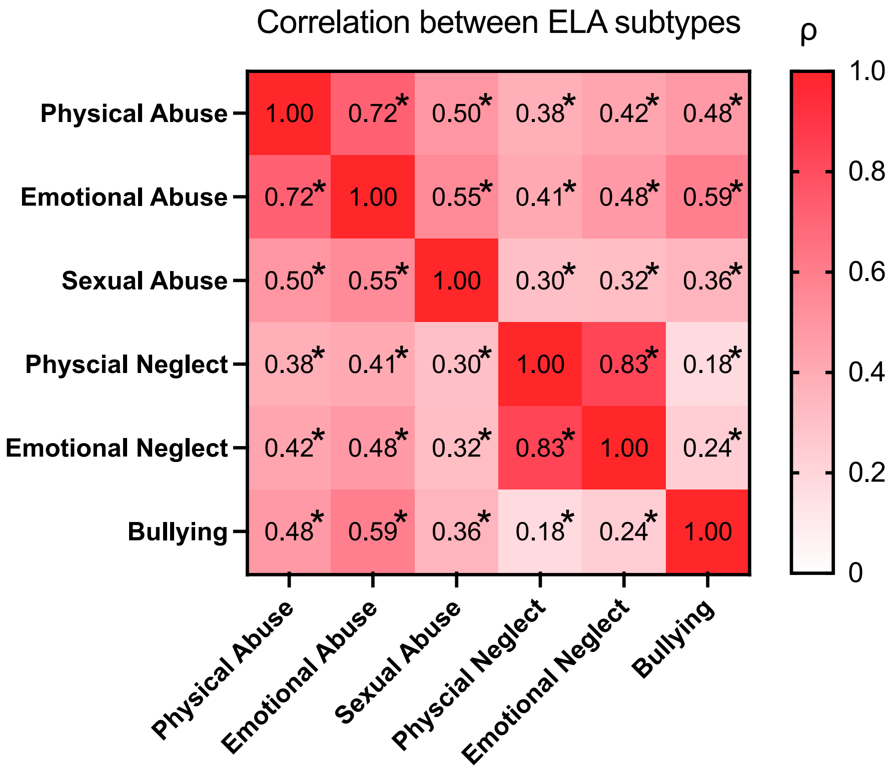


**Supplementary Figure 1.** Correlation matrix of each subtype of ELA and childhood bullying. Spearman's ρ is displayed in each box. Each subtype was statistically significantly correlated with every other subtype, with all *p*-values<0.001. The most highly correlated pairs of ELA subtypes were physical and emotional neglect (Spearman's ρ=0.83, *p*<0.001) and physical and emotional abuse (Spearman's ρ=0.72, *p*<0.001).

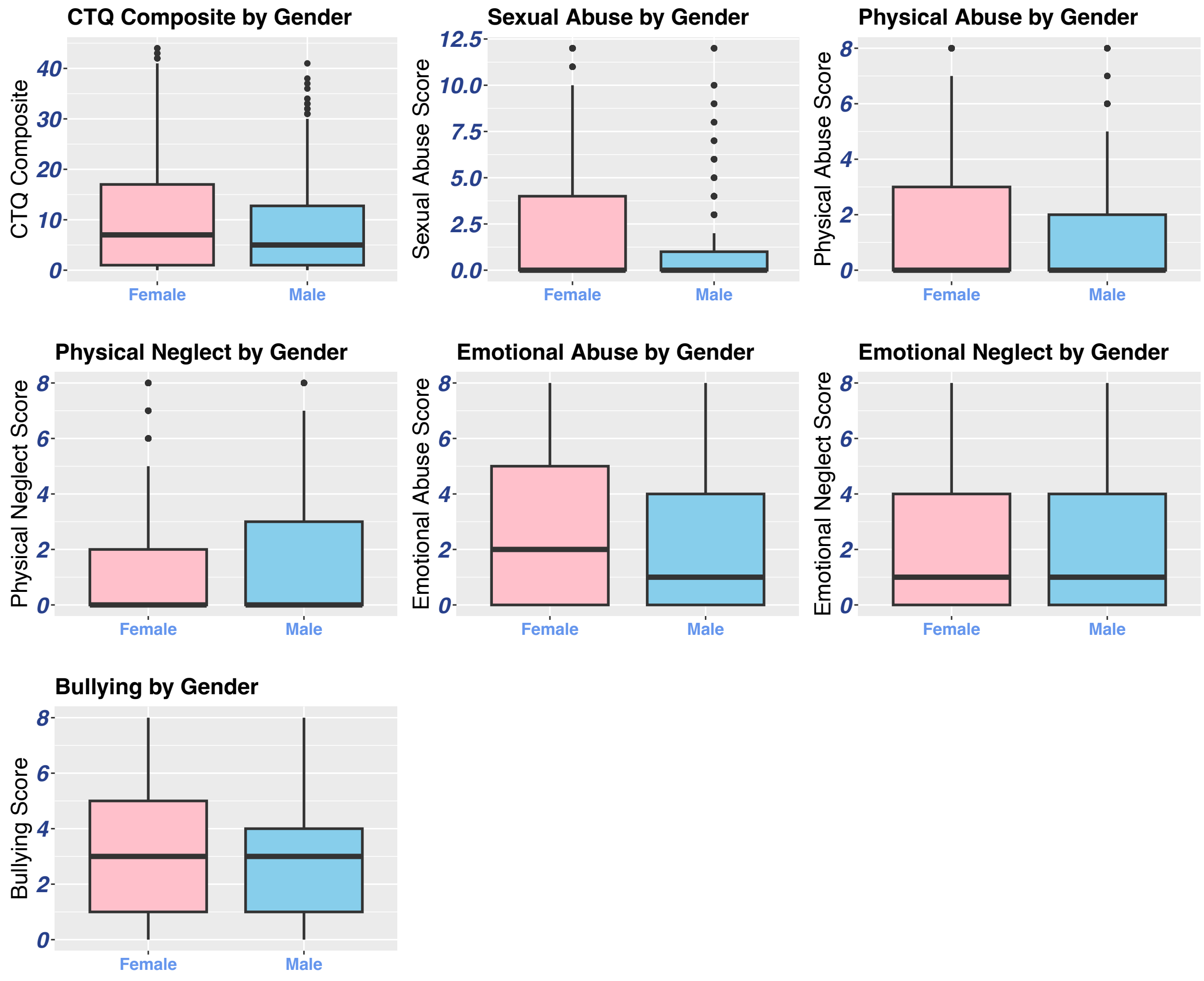


**Supplementary Figure 2. Sex differences between CTQ composite scores and each ELA subtype.** Each bar graph shows the mean and standard deviation of each ELA subscore, stratified by male and female gender on birth certificate. Welch's test was used for each comparison. CTQ composite score, sexual abuse, physical abuse, and emotional abuse scores were significantly greater in women than men (CTQ: t(2265.3) = 5.9715, $p<0.001$; sexual abuse: t(2462.5)=10.87, $p<0.001$; physical abuse: t(2199.9)= 3.86, $p<0.001$; emotional abuse: t(2232.8)=8.05, $p<0.001$). Physical neglect was significantly greater in men (t(1876.9)= -2.0304, $p=0.042$). Bullying and emotional neglect were not significantly different across men and women ($p>0.05$).

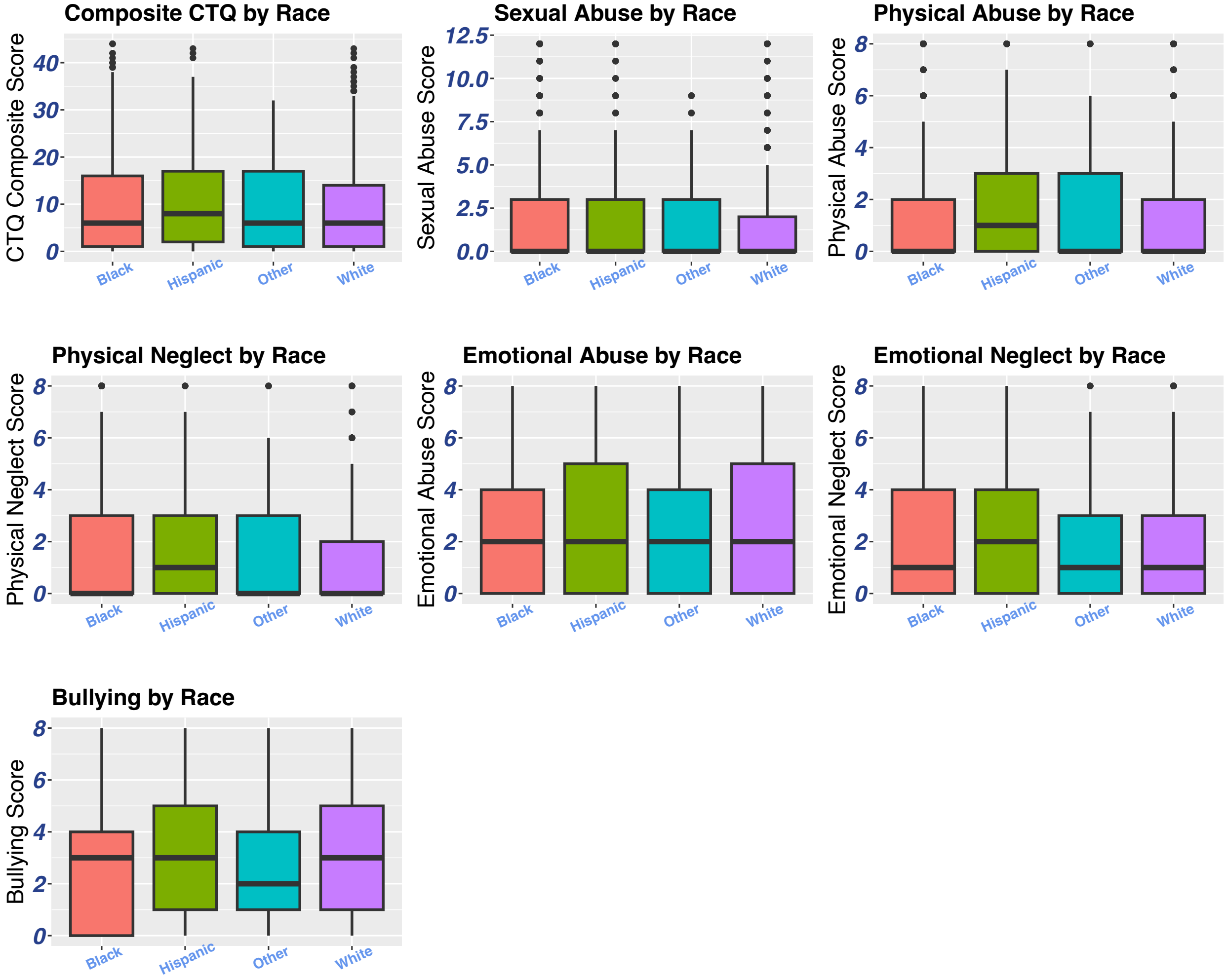


**Supplementary Figure 3. Racial/ethnic differences between CTQ composite scores and each ELA subtype.** Each bar graph shows the mean and standard deviation of each ELA subscore, stratified by self-reported race/ethnicity. CTQ composite score differed marginally by race/ethnicity groups (Kruskal-Wallis $\chi2(3)=7.79$, $p=0.051$) with Hispanic individuals showing slightly higher rates of ELA compared to non-hispanic white, non-hispanic black, and other groups.

**Supplementary Table 1**. Distribution of socio-demographic, ED/trauma-related, and past pain/stress variables based on CPMP trajectory

| | Low Pain (n=1070) | | Moderate Recovery (n=468) | | Moderate Pain (n=590) | | High Pain (n=352) | | $\chi^2$ | *p*-value |
|---|---|---|---|---|---|---|---|---|---|---|
| | **% or Mean** | **(n) or (SD)** | **% or Mean** | **(n) or (SD)** | **% or Mean** | **(n) or (SD)** | **% or Mean** | **(n) or (SD)** | | |
| **Socio-demographic variables** | | | | | | | | | | |
| Female | 57% | 605 | 65% | 303 | 69% | 409 | 67% | 237 | 32.46 | <0.001 |
| Age | 33.36 | 12.52 | 36.81 | 14.00 | 37.61 | 13.12 | 40.89 | 13.19 | 109.34 | <0.001 |
| Race/ethnicity | | | | | | | | | 36.86 | <0.001 |
| Hispanic | 11% | 122 | 9% | 42 | 13% | 77 | 11% | 37 | | |
| Non-Hispanic white | 36% | 387 | 45% | 210 | 34% | 201 | 26% | 93 | | |
| Non-Hispanic Black | 49% | 521 | 42% | 197 | 49% | 285 | 60% | 210 | | |
| Other | 4% | 39 | 4% | 17 | 390% | 23 | 3% | 12 | | |
| BMI | 29.17 | 7.95 | 29.75 | 7.93 | 30.95 | 8.28 | 32.15 | 9.44 | 39.85 | <0.001 |
| Area Deprivation Index | 62.03 | 28.04 | 60.04 | 27.52 | 66.28 | 26.76 | 70.74 | 26.08 | 37.07 | <0.001 |
| **ED/Trauma-related variables** | | | | | | | | | | |
| Site | | | | | | | | | 7.12 | 0.310 |
| Northeast | 41% | 441 | 45% | 209 | 45% | 267 | 39% | 136 | | |
| Southeast | 18% | 197 | 18% | 86 | 18% | 107 | 22% | 77 | | |
| Midwest | 40% | 432 | 37% | 173 | 37% | 216 | 39% | 139 | | |
| Trauma Type | | | | | | | | | 10.05 | 0.123 |
| MVC | 76% | 814 | 76% | 358 | 79% | 469 | 76% | 268 | | |
| Physicalsexual assault | 10% | 104 | 8% | 38 | 8% | 45 | 13% | 44 | | |
| Other | 14% | 152 | 15% | 72 | 13% | 76 | 11% | 40 | | |
| Peritraumatic Distress Inventory | 12.49 | 7.09 | 14.49 | 7.13 | 15.00 | 7.07 | 15.81 | 7.33 | 74.36 | <0.001 |
| **Past pain/stress variables** | | | | | | | | | | |
| Had previous moderate/severe pain | 18% | 189 | 27% | 123 | 43% | 251 | 61% | 214 | 275.62 | <0.001 |
| Childhood Trauma Questionnaire | | | | | | | | | | |
| Any Childhood Trauma (CTQ≥1) | 75% | 805 | 81% | 381 | 83% | 490 | 86% | 301 | 25.50 | <0.001 |
| CTQ Composite Score (0-44) | 8.02 | 9.02 | 9.37 | 9.54 | 10.75 | 10.33 | 12.09 | 10.63 | | |
| Any Physical Abuse | 37% | 397 | 43% | 201 | 51% | 298 | 55% | 193 | 47.78 | <0.001 |
| Physical Abuse Subscore (0-8) | 1.23 | 2.08 | 1.38 | 2.13 | 1.85 | 2.41 | 2.3 | 2.74 | 64.80 | <0.001 |
| Any Emotional Abuse | 58% | 623 | 70% | 328 | 71% | 416 | 74% | 260 | 46.92 | <0.001 |
| Emotional Abuse Subscore (0-8) | 2.07 | 2.45 | 2.55 | 2.51 | 2.94 | 2.71 | 3.4 | 2.92 | 80.57 | <0.001 |
| Any Sexual Abuse | 28% | 297 | 39% | 184 | 40% | 238 | 42% | 148 | 43.60 | <0.001 |
| Sexual Abuse Susbcore (0-12) | 1.43 | 2.94 | 2 | 3.24 | 2.37 | 3.65 | 2.63 | 3.87 | 49.96 | <0.001 |
| Any Physical Neglect | 43% | 460 | 45% | 212 | 50% | 297 | 50% | 177 | 11.01 | 0.012 |
| Physcial Neglect Subscore (0-8) | 1.47 | 2.22 | 1.44 | 2.03 | 1.61 | 2.08 | 1.69 | 2.17 | 9.13 | 0.028 |
| Any Emotional Neglect | 49% | 525 | 52% | 243 | 55% | 327 | 54% | 189 | 6.81 | 0.078 |
| Emotional Neglect Subscore (0-8) | 1.81 | 2.36 | 2 | 2.39 | 1.98 | 2.27 | 2.07 | 2.43 | 6.76 | 0.080 |
| Bullying | | | | | | | | | | |
| Any | 76% | 814 | 85% | 397 | 81% | 476 | 82% | 290 | 18.23 | <0.001 |
| Bullying Score (0-8) | 2.60 | 2.25 | 3.05 | 2.25 | 3.17 | 2.42 | 3.63 | 2.63 | 52.79 | <0.001 |

ED-emergency department; CPMP-Chronic Posttraumatic Musculoskeletal Pain; SD-standard deviation; BMI-body mass index; MVC, motor vehicle collision;
*Significant at the 0.05 level, two-sided test

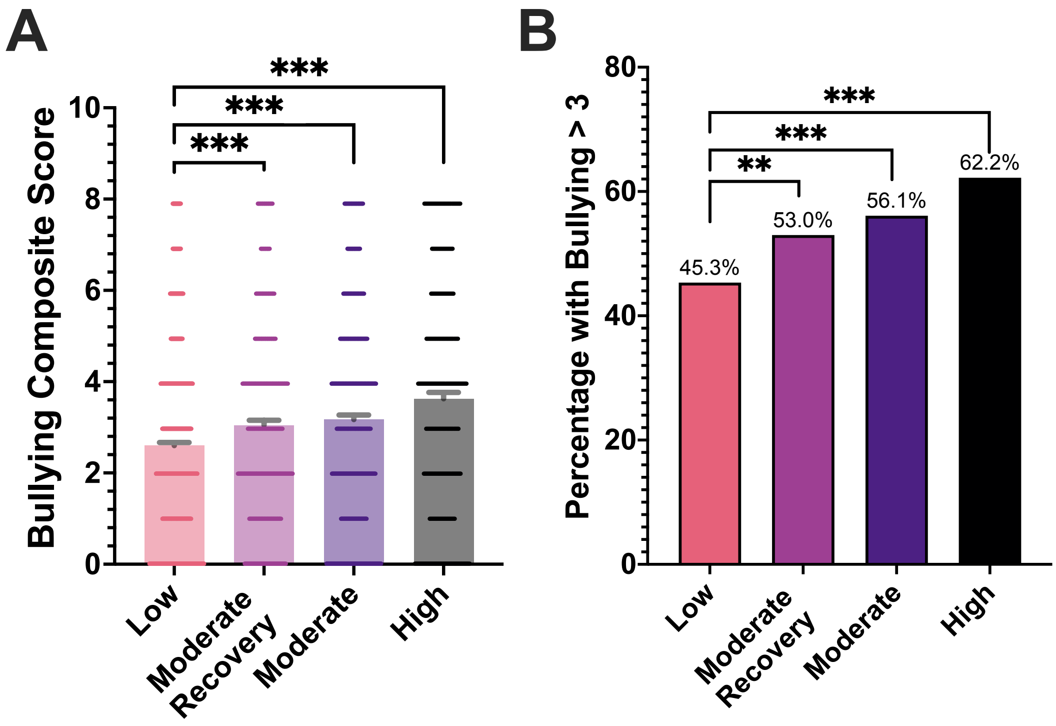


**Supplementary Figure 4.** The relationship between the four latent class groups and bullying score. **A**) Average bullying scores in the four latent class groups and **B)** percentage of individuals with a bullying score above the median (bullying>3) in each latent class group are shown. Bars show mean and error bars show standard error of the mean. Individual values are represented by dots overlaid onto the bar charts. *Significance: **p<0.01, ***p<0.001*

**Supplementary Table 2.** Relative Risk Statistics

| | Exposure to Past Trauma | Prevalence of > 0 or high (in 2 groups) | Prevalence of Outcome* (in 2 groups) | Relative Risk (RR) | P Value* | Population Attributable Risk (PAR)* | Population Attributable Fraction (PAF)* |
|---|---|---|---|---|---|---|---|
| **Low Pain vs. Moderate Pain Trajectory** | ELA (none vs. high) | 60% | 37% | 1.58 | **<0.001** | 0.09 | 26% |
| | ELA (none vs. any) | 78% | 36% | 1.38 | **<0.001** | 0.08 | 23% |
| | Emotional Abuse (none vs. any) | 63% | 36% | 1.43 | **<0.001** | 0.08 | 21% |
| | Physical Abuse (none vs. any) | 42% | 36% | 1.42 | **<0.001** | 0.05 | 15% |
| | Sexual Abuse (none vs. any) | 32% | 36% | 1.42 | **<0.001** | 0.04 | 12% |
| | Physical Neglect (none vs. any) | 46% | 36% | 1.21 | **0.005** | 0.03 | 9% |
| | Emotional Neglect (none vs. any) | 51% | 36% | 1.18 | **0.015** | 0.03 | 8% |
| **Low Pain vs. Moderate Recovery Trajectory** | ELA (none vs. high) | 57% | 29% | 1.33 | **0.013** | 0.05 | 16% |
| | ELA (none vs. any) | 77% | 30% | 1.30 | **0.010** | 0.06 | 19% |
| | Emotional Abuse (none vs. any) | 62% | 30% | 1.45 | **<0.001** | 0.07 | 22% |
| | Physical Abuse (none vs. any) | 39% | 30% | 1.18 | **0.035** | 0.02 | 7% |
| | Sexual Abuse (none vs. any) | 31% | 30% | 1.42 | **<0.001** | 0.04 | 12% |
| | Physical Neglect (none vs. any) | 44% | 30% | 1.07 | 0.433 | 0.01 | 3% |
| | Emotional Neglect (none vs. any) | 50% | 30% | 1.08 | 0.329 | 0.01 | 4% |
| **Low Pain vs. High Pain Trajectory** | ELA (none vs. high) | 60% | 27% | 2.13 | **<0.001** | 0.11 | 40% |
| | ELA (none vs. any) | 78% | 25% | 1.69 | **<0.001** | 0.09 | 35% |
| | Emotional Abuse (none vs. any) | 62% | 25% | 1.73 | **<0.001** | 0.08 | 31% |
| | Physical Abuse (none vs. any) | 41% | 25% | 1.71 | **<0.001** | 0.06 | 23% |
| | Sexual Abuse (none vs. any) | 31% | 25% | 1.59 | **<0.001** | 0.04 | 16% |
| | Physical Neglect (none vs. any) | 45% | 25% | 1.25 | **0.020** | 0.02 | 10% |
| | Emotional Neglect (none vs. any) | 50% | 25% | 1.15 | 0.148 | 0.02 | 7% |
| **Moderate Pain vs. High Pain Trajectory** | ELA (none vs. high) | 72% | 39% | 1.21 | 0.158 | 0.05 | 13% |
| | ELA (none vs. any) | 84% | 37% | 1.13 | 0.366 | 0.04 | 10% |
| | Emotional Abuse (none vs. any) | 72% | 37% | 1.11 | 0.302 | 0.03 | 7% |
| | Physical Abuse (none vs. any) | 52% | 37% | 1.11 | 0.224 | 0.02 | 6% |
| | Sexual Abuse (none vs. any) | 41% | 37% | 1.05 | 0.655 | 0.01 | 2% |
| | Physical Neglect (none vs. any) | 50% | 37% | 1.00 | 1.000 | 0.00 | 0% |
| | Emotional Neglect (none vs. any) | 55% | 37% | 0.96 | 0.654 | -0.01 | -2% |
| **Moderate Recovery vs. High Pain Trajectory** | ELA (none vs. high) | 69% | 47% | 1.40 | **0.006** | 0.10 | 22% |
| | ELA (none vs. any) | 83% | 43% | 1.19 | 0.144 | 0.06 | 14% |
| | Emotional Abuse (none vs. any) | 72% | 43% | 1.12 | 0.267 | 0.03 | 8% |
| | Physical Abuse (none vs. any) | 48% | 43% | 1.31 | **0.001** | 0.06 | 13% |
| | Sexual Abuse (none vs. any) | 40% | 43% | 1.07 | 0.474 | 0.01 | 3% |
| | Physical Neglect (none vs. any) | 47% | 43% | 1.12 | 0.179 | 0.02 | 5% |
| | Emotional Neglect (none vs. any) | 53% | 43% | 1.04 | 0.666 | 0.01 | 2% |

**Prevalence of Outcome: Prevalence of the higher pain trajectory class in the two classes of comparison*

**P Value: from Chisquare test between outcome and 2 groups of Past Trauma*

**PAR = P(Pain in population) - P(Pain in 0, or no, or low)*

**PAF = PAR /P(Pain in population)*

*ELA-early life adversity; RR-relative risk; PAR- population attributable risk; PAF-population attributable fraction*

**Supplementary Table 3**. Model Summary with ED Opioid Adminstration Covariate

| **low pain group as reference* | | Complete Model (n=2480) | | | + Opioids in the ED (n=1,991) | | |
|---|---|---|---|---|---|---|---|
| | | OR | (95% CI) | *BH-adj. p* | OR | (95% CI) | *BH-adj. p* |
| **High Pain** | CTQ | 1.030 | 1.016 - 1.045 | **<0.001** | 1.033 | 1.017 - 1.049 | **<0.001** |
| | Opioids in the ED | | | | 1.378 | 0.977 - 1.944 | 0.135 |
| **Moderate Pain** | CTQ | 1.022 | 1.010 - 1.035 | **0.001** | 1.024 | 1.011 - 1.038 | **<0.001** |
| | Opioids in the ED | | | | 1.260 | 0.951 - 1.670 | 0.201 |
| **Moderate Recovery** | CTQ | 1.013 | 1.000 - 1.026 | 0.119 | 1.019 | 1.005 - 1.034 | **0.021** |
| | Opioids in the ED | | | | 1.479 | 1.108 - 1.974 | **0.021** |

*Each model was also adjusted for Age, ED site, race/ethnicity, sex,marital status, BMI, previous pain, area deprivation index, peritraumatic distress inventory, and trauma type*

*ED-emergency department; CTQ-childhood trauma questionnaire score; OR-odds ratio; CI-confidence interval; BH-Benjamini Hochberg; BMI-body mass index*

Significant *p*-values are shown in bold

**Supplementary Table 4**. Summary Statistics for Z-score Normalized ELA Subscores and CPMP Outcome

| ** low pain as reference group* | Moderate Recovery | | | | Moderate Pain | | | | High Pain | | | |
|---|---|---|---|---|---|---|---|---|---|---|---|---|
| **ELA Subtype** | OR | 95% CI | *p* | *BH-adj. p* | OR | 95% CI | *p* | *BH-adj. p* | OR | 95% CI | *p* | *BH-adj. p* |
| Physical abuse | | | | | | | | | | | | |
| CTQ subscore | 1.028 | 0.902 - 1.171 | 0.680 | 0.982 | 1.251 | 1.116 - 1.404 | **<0.001** | **0.003** | 1.382 | 1.207 - 1.582 | **<0.001** | **<0.001** |
| Sexual abuse | | | | | | | | | | | | |
| CTQ subscore | 1.153 | 1.012 - 1.313 | **0.032** | 0.230 | 1.203 | 1.068 - 1.355 | **0.002** | 0.053 | 1.202 | 1.043 - 1.385 | **0.011** | 0.131 |
| Emotional abuse | | | | | | | | | | | | |
| CTQ subscore | 1.154 | 1.016 - 1.312 | **0.028** | 0.230 | 1.317 | 1.168 - 1.485 | **<0.001** | **<0.001** | 1.502 | 1.299 - 1.738 | **<0.001** | **<0.001** |
| Physical neglect | | | | | | | | | | | | |
| CTQ subscore | 1.028 | 0.909 - 1.163 | 0.656 | 0.982 | 1.060 | 0.945 - 1.190 | 0.321 | 0.834 | 1.103 | 0.958 - 1.269 | 0.172 | 0.632 |
| Emotional neglect | | | | | | | | | | | | |
| CTQ subscore | 1.125 | 0.999 - 1.268 | 0.052 | 0.385 | 1.052 | 0.937 - 1.181 | 0.394 | 0.834 | 1.106 | 0.961 - 1.273 | 0.158 | 0.600 |
| Bullying | | | | | | | | | | | | |
| Subscore | 1.161 | 1.024 - 1.317 | **0.020** | 0.191 | 1.234 | 1.095 - 1.390 | **0.001** | **0.011** | 1.425 | 1.232 - 1.648 | **<0.001** | **<0.001** |

*ELA-early life adversity; OR-odds ratio; CI-confidence interval; BH-Benjamini Hochberg; CTQ-childhood trauma questionnaire score*

Significant *p* -values are shown in bold
